# National and Subnational Level Estimates of Maternal Delivery at Home and Their Predictors in Bangladesh: Evidence from Population-Based Survey 2012-2022

**DOI:** 10.1101/2025.07.05.25330941

**Authors:** Raisha Binte Islam, Md. Lutful Kader, Somaya Mostarin, Mushfika Binta Latif, Amina Anjum, Syed Toukir Ahmed Noor

**Author notes:** Corresponding author Syed Toukir Ahmed Noor Department of Statistics, Shahjalal University of Science and Technology, Sylhet-3114, Bangladesh.

## Abstract

**Background:** Despite significant progress in reducing maternal and neonatal mortality, home delivery remains a substantial public health challenge in Bangladesh and many other low- and middle-income countries. While the proportion of home deliveries has markedly decreased in the past decade, pronounced disparities persist across geographic, socioeconomic, and demographic groups. A nuanced understanding of the prevalence and determinants of maternal home delivery is key to designing targeted interventions. This study examines national and subnational variations in maternal home delivery and associated factors in Bangladesh.

**Methods:** We analyzed data from the Multiple Indicator Cluster Surveys (2012–13 and 2019) and the Bangladesh Demographic and Health Survey (2022), covering 20,770 ever-married women aged 15–49 who gave birth in the preceding two years. District-level prevalence, descriptive statistics, and multivariable logistic regression were used to assess trends and determinants.

**Results:** Home delivery prevalence declined from 68% in 2012–13 to 35% in 2022. Disparities remain: divisions such as Barisal (48.9%), Chattogram, Sylhet, and Mymensingh showed higher rates, while Dhaka and Khulna had the lowest. At the subnational level, remote areas like Bandarban, Rangamati, and Bhola exhibited higher prevalence. The logistic regression analysis identified several significant predictors, such as women with no formal education, limited ANC visits (≤3), rural residence, lower wealth status, multiparity (≥3 children), and lack of media exposure were more likely to deliver at home.

**Conclusions:** Despite marked improvement, persistent geographic and socioeconomic inequities highlight the need for targeted interventions. Strengthening healthcare infrastructure in underserved regions, promoting maternal health awareness, scaling up ANC utilization, and reducing financial barriers through subsidies and incentive programs can help further decrease home deliveries. Future research should explore cultural and religious factors to inform context-specific policies for equitable facility-based childbirth.

## Introduction

Despite a 40% reduction in the global maternal mortality ratio (MMR)-from 328 deaths per 100,000 live births in 2000 to 197 in 2023-over 90% of all maternal deaths continue to occur in low- and lower-middle-income countries [1]. South Asia remains a critical contributor, accounting for nearly one-quarter of global maternal fatalities [2]. As a prominent example, Bangladesh, a lower-middle-income country in South Asia, has made commendable progress in reducing its MMR from 412 deaths per 100,000 live births in 1976 to 123 in 2020 [3]. This progress has been accompanied by significant scale-up in key maternal health interventions, including antenatal care (4+ visits) from 17% in 2004 to 41% in 2022, facility-based delivery from 10% in 2004 to 65% in 2022, skilled birth attendance from 23% in 2007 to 70% in 2022, and postnatal care (by medically trained providers within 48 hours) from 16% in 2004 to 55% in 2022 [4]. Despite these advances, the gains have been unevenly distributed across geographic regions and socioeconomic strata, resulting in persistent disparities in maternal health outcomes. Consequently, Bangladesh remains off track to meet the Sustainable Development Goal target (SDG 3.1), which targets an MMR of fewer than 70 deaths per 100,000 live births by 2030 [5].

To accelerate the progress toward this ambitious goal, there is an urgent need to improve emergency obstetric services and minimize the incidence of maternal delivery at home, which are associated with preventable maternal and neonatal mortality. Globally, maternal delivery at home accounted for an estimated 287,000 maternal and 2.4 million newborn deaths in 2020 alone [6,7]. A study from Nigeria found that non-institutional delivery (delivery outside health facilities, often by unskilled attendants) was associated with a 79.2% chance of maternal death among the cases studied [8]. In Bangladesh, more than one-third of all deliveries still occur at home, with marked regional disparities across administrative divisions [4] Reducing home deliveries is, therefore, a critical public health priority to advance equity and reduce mortality and neonatal mortality [9–11]. Maternal delivery at home is influenced by a complex interplay of sociodemographic, economic, cultural, and health system factors. Key factors include maternal education, quantity of antenatal care visits, parity, household wealth, religion, and geographic accessibility to healthcare facilities [12–14]. In addition, entrenched poverty, traditional and religious norms, poor transportation infrastructure, and the limited autonomy among women in decision-making further exacerbate the preference for delivery at home [15]. Other factors such as low maternal and paternal education, advanced maternal age, multiparity, and low wealth status also contribute to increased home delivery practices [16]. Fear of cesarean sections, perceptions of poor quality of care, and the lack of female doctors also play substantial roles in influencing delivery preferences [15].

According to the availability, accessibility, acceptability, and quality (AAAQ) framework, both supply-side (e.g. healthcare infrastructure and provider availability) and demand-side (e.g. household-level barriers and perceptions) factors must be addressed to reduce maternal delivery at home [17]. While individual and household-level determinants have been widely studied, health system constraints-especially at the subnational level-remain largely underexplored in the context of Bangladesh. Identifying subnational areas (here district in Bangladesh) with persistently high rates of maternal delivery at home is essential for designing and implementing evidence-based maternal health interventions.

However, limited research has assessed the sub-national level distribution of maternal delivery at home in Bangladesh [18–20]. Moreover, little is known about how associated factors of maternal delivery at home vary over time across urban and rural settings. To our knowledge, no prior study has investigated subnational variations in maternal delivery at home and explored the evolving determinants of this practice. Therefore, we aimed to investigate national and subnational patterns of maternal delivery at home and analyze the associated factors as a whole as well as in urban and rural areas in Bangladesh. The findings will inform targeted, context-specific policies and support the implementation of effective maternal health interventions.

## Materials and Methods

### Data Sources

This study utilized secondary data derived from three nationally representative, population-based cross-sectional surveys: the Multiple Indicator Cluster Survey (MICS) conducted in 2012–13 and 2019, and the Bangladesh Demographic and Health Survey (BDHS) conducted in 2022. The MICS, implemented by the Bangladesh Bureau of Statistics (BBS) with technical and financial support from the United Nations International Children’s Emergency Fund (UNICEF), provides comprehensive data on household characteristics and maternal and child health indicators at the district level. The BDHS, administered under the authority of the National Institute of Population Research and Training (NIPORT), Medical Education and Family Welfare Division, Ministry of Health and Family Welfare (MOHFW), offers vital demographic and health statistics, including district-level estimates for the first time in 2022.

### Sampling Design and Sample Size

Both the MICS and BDHS employed a uniform methodology to ensure national representativeness. Detailed methodological descriptions are available in the respective technical reports [4,21,22]. Briefly, the MICS applied a multistage stratified cluster sampling strategy, covering all 64 administrative districts of Bangladesh. Each district was stratified into urban and rural areas. Enumeration areas (EAs) were selected using the probability proportional to size (PPS) method within these strata. From each selected EA, 20 households were systematically sampled [23,24]. The MICS sampled a total of 55,120 in 2012-2013 and 64,400 households in 2019, of which 51,895 and 61,242 were successfully interviewed, with response rates of 98.5% and 99.4%, respectively. After excluding respondents with no delivery history or missing birth outcome data, the final analytical sample included 7795 and 9284 women aged 15-49 years with a history of childbirth in 2012-2013 and 2019, respectively.

The BDHS 2022 utilized a two-stage stratified sampling strategy based on the 2011 Population Census. In the first stage, 675 EAs were selected (237 urban and 438 rural) using PPS sampling. In the second stage, 45 households were systematically selected from each EA to ensure adequate representation across residence types, and for the first time, BDHS provided the subnational level variable, which offers an opportunity to analyze the subnational level estimate [25]. In 2022, the final response rate of BDHS was 99.6%, with 30,330 households selected and 30,018 households interviewed. The final analytical sample from BDHS 2022 consisted of 3,691 women of reproductive age (15–49 years) with recent birth histories. We conducted a complete case analysis; as a result, we omitted all missing observations in our study. Women aged 15-49 with a live birth in the past 2 years were considered for all the rounds.

### Outcome Variable

The main outcome variable for this study was maternal delivery at home, defined dichotomously. Deliveries occurring at the respondent’s home were coded as “1” (maternal delivery at home), and those occurring at any health facility were coded as “0”.

### Explanatory Variables

A total of 13 independent variables were selected on an extensive review of previous literature [13,26,27], capturing individual-, household-, and community-level determinants. Individual-level factors included women’s age, education (no education, primary, secondary, and higher), number of antenatal care (ANC) visits (none, 1, 2, 3, and ≥4), parity (1–2, ≥3), media exposure, religion (Non-Muslim, Muslim), husband’s age, and spousal age gap (≤5 years, 6–9 years, and more than 9 years). Household-level factors included education of household’s head, household size and household’s socioeconomic status. The community-level factors encompassed administrative division and place of residence (urban or rural). Socioeconomic status was assessed using the wealth index, constructed via principal component analysis (PCA) as provided in MICS and BDHS datasets. Households were classified into five quintiles: poorest, poorer, middle, richer, and richest [4,21,22].

### Statistical analysis

The data were analyzed via Stata version 17.0 (StataCorp, College Station, Texas, USA), whereas data visualization was performed via R version 4.3.0. To account for the complex survey design, we incorporated primary sampling units (PSUs) and strata and applied sampling weights to ensure representativeness. The Stata “svyset” command was used to adjust for the survey design [28], and all analyses adhered to the STROBE cross-sectional reporting guidelines [29].

Descriptive statistics were used to summarize the characteristics of the study population, which were reported as frequencies and proportions in percentages. We conducted bivariate analysis using the Chi-square test to assess associations between the explanatory variables and the outcome variable. We employed simple binary logistic regression analysis to estimate crude odds ratios (ORs) with 95% CIs for each explanatory variable in the unadjusted analysis. Variables that were statistically significant in the simple logistic regression models were subsequently included in the multiple logistic regression model to estimate adjusted odds ratios (AORs) with their respective 95% CIs. Statistical significance was determined at the 5% level (p-value <0.05). Multicollinearity was checked for all the variables through VIF, and the value of VIF was <5 for all variables.

To account for the complex survey design, we applied the “svyset” command in Stata, incorporating sampling weights, strata, and primary sampling units (PSUs). All the analyses were done using the “svy” prefix to ensure design-based estimates and inference [28]. The study adhered to the STROBE guidelines for reporting cross-sectional studies [29]. We used Stata version 17.0 (StataCorp, College Station, Texas, USA) for data analysis, R version 4.3.0 for data visualization and ArcGIS version 10.8 for mapping.

### Ethical Consideration

This study utilized deidentified secondary data obtained from the UNICEF (https://mics.unicef.org/surveys) and DHS (https://www.dhsprogram.com/data/available-datasets.cfm) websites. Since the data were publicly available and did not contain any personally identifiable information, ethical approval was not needed. All participants provided informed consent, and the original surveys were carried out in accordance with ethical standards and approved by the appropriate institutional review boards.

## Results

### Sample characteristics

Across all survey years, approximately one-third of women were aged 20-24 years (33.6% in 2012-13, 32.2% in 2019, and 33.8% in 2022). Educational attainment demonstrated a positive trend over time, the proportion of women completing higher education increased from 12.4% in 2012 to 18.5% in 2022. Nearly nine in 10 women were Muslim (89.2% in 2012, 90.4% in 2019 and 92.7% in 2022). Media exposure declined slightly over the years, from 76.6% in 2012 to 66.3% in 2022. Utilization of ANC services improved, with the proportion of women receiving at least four ANC visits increased from 22.7% in 2012 to 40.5% in 2022. Fertility pattern showed that majority of women had 1–2 children, increasing from 65.6% in 2012 to 71.9% in 2022. The proportion of households headed by individuals with no education accounted for 56.2% in 2012, 30.3% in 2019 and 15.5% in 2022. The proportion of women in the poorest wealth quintile declined from 28.3% in 2012 to 20.2% in 2022. Household size remained relatively stable, with approximately two-thirds of women living in households comprising five or more members (65.6% in 2012-13, 63% in 2019 and 69.5% in 2022). Most women resided in rural areas (83.8% in 2012, 80.9% in 2019 and 73% in 2022), and the Dhaka division consistently had the highest representation across survey years (**Table 1**).

**Table 1:** Sample characteristics of the respondents, presented in terms of weighted percentage.

| Variables | 2012-13 |  | 2019 |  | 2022 |  |
| --- | --- | --- | --- | --- | --- | --- |
|  | Number | % | Number | % | Number | % |
| ***Age of women (in years)*** |  |  |  |  |  |  |
| 15-19 | 906 | 11.6 | 1,234 | 13.3 | 614 | 16.6 |
| 20-24 | 2,615 | 33.6 | 2,987 | 32.2 | 1248 | 33.8 |
| 25-29 | 2,351 | 30.2 | 2,562 | 27.6 | 962 | 26.1 |
| 30-34 | 1,143 | 14.7 | 1,640 | 17.7 | 592 | 16.0 |
| 35-49 | 780 | 10.0 | 861 | 9.3 | 274 | 7.4 |
| ***Education of women*** |  |  |  |  |  |  |
| No education | 1,621 | 20.8 | 846 | 9.1 | 183 | 5.0 |
| Primary | 2,294 | 29.4 | 2,151 | 23.2 | 831 | 22.5 |
| Secondary | 2,917 | 37.4 | 4,691 | 50.5 | 1995 | 54.0 |
| Higher | 963 | 12.4 | 1,596 | 17.2 | 682 | 18.5 |
| **Religion** |  |  |  |  |  |  |
| Non-Muslim | 842 | 10.8 | 896 | 9.7 | 269 | 7.3 |
| Muslim | 6,953 | 89.2 | 8,388 | 90.4 | 3422 | 92.7 |
| **Media exposure** |  |  |  |  |  |  |
| No | 1,186 | 23.5 | 3,308 | 35.7 | 1243 | 33.7 |
| Yes | 3,872 | 76.6 | 5,967 | 64.3 | 2447 | 66.3 |
| **Number of ANC visits** |  |  |  |  |  |  |
| No visit | 2,730 | 35.2 | 1,677 | 18.1 | 268 | 7.4 |
| 1 visit | 1,018 | 13.1 | 1,123 | 12.1 | 526 | 14.6 |
| 2 visits | 1,227 | 15.8 | 1,611 | 17.4 | 694 | 19.2 |
| 3 visits | 1,024 | 13.2 | 1,529 | 16.5 | 658 | 18.2 |
| 4+ visits | 1,758 | 22.7 | 3,339 | 36.0 | 1462 | 40.5 |
| **Parity** |  |  |  |  |  |  |
| 1-2 | 5,116 | 65.6 | 6,401 | 69.0 | 2653 | 71.9 |
| 3+ | 2,679 | 34.4 | 2,883 | 31.1 | 1037 | 28.1 |
| **Age of husband (in year)** |  |  |  |  |  |  |
| 15-29 | 2,514 | 32.9 | 2,789 | 30.3 | 1237 | 33.8 |
| 30-34 | 1,960 | 25.6 | 2,627 | 28.5 | 913 | 24.9 |
| 35-39 | 1,606 | 21.0 | 2,062 | 22.4 | 832 | 22.7 |
| 40-44 | 872 | 11.4 | 988 | 10.7 | 407 | 11.1 |
| 45-49 | 409 | 5.4 | 505 | 5.5 | 191 | 5.2 |
| 50-90 | 290 | 3.8 | 243 | 2.6 | 79 | 2.2 |
| **Spousal age gap (in year)** |  |  |  |  |  |  |
| ≤5 | 2,939 | 38.0 | 3,534 | 38.4 | 1408 | 38.5 |
| 6-9 | 2,630 | 34.0 | 3,088 | 33.5 | 1192 | 32.6 |
| >9 | 2,173 | 28.1 | 2,593 | 28.1 | 1060 | 29.0 |
| **Education of household head** |  |  |  |  |  |  |
| No education | 4,375 | 56.2 | 2,812 | 30.3 | 566 | 15.5 |
| Primary | 1,009 | 13.0 | 2,794 | 30.1 | 1059 | 28.9 |
| Secondary | 1,410 | 18.1 | 2,602 | 28.0 | 1277 | 34.9 |
| Higher | 992 | 12.7 | 1,073 | 11.6 | 757 | 20.7 |
| **Wealth index** |  |  |  |  |  |  |
| Richest | 1,174 | 15.1 | 1,587 | 17.1 | 661 | 17.9 |
| Fourth | 1,283 | 16.5 | 1,770 | 19.1 | 718 | 19.5 |
| Middle | 1,474 | 18.9 | 1,810 | 19.5 | 787 | 21.3 |
| Second | 1,661 | 21.3 | 1,886 | 20.3 | 780 | 21.1 |
| Poorest | 2,203 | 28.3 | 2,231 | 24.0 | 745 | 20.2 |
| **Household size** |  |  |  |  |  |  |
| ≤4 | 2684 | 34.4 | 3,439 | 37.0 | 1128 | 30.6 |
| 5+ | 5,111 | 65.6 | 5,845 | 63.0 | 2563 | 69.5 |
| **Residence** |  |  |  |  |  |  |
| Urban | 1,261 | 16.2 | 1,774 | 19.1 | 997 | 27.0 |
| Rural | 6,534 | 83.8 | 7,510 | 80.9 | 2694 | 73.0 |
| **Division** |  |  |  |  |  |  |
| Barisal | 703 | 9.0 | 835 | 9.0 | 219 | 5.9 |
| Chittagong | 1,585 | 20.3 | 1,926 | 20.8 | 793 | 21.5 |
| Dhaka | 2,002 | 25.7 | 1,795 | 19.3 | 926 | 25.1 |
| Khulna | 1,044 | 13.4 | 1,275 | 13.7 | 380 | 10.3 |
| Mymensingh |  |  | 558 | 6.0 | 333 | 9.0 |
| Rajshahi | 715 | 9.2 | 956 | 10.3 | 381 | 10.3 |
| Rangpur | 972 | 12.5 | 1,135 | 12.2 | 419 | 11.4 |
| Sylhet | 774 | 9.9 | 804 | 8.7 | 240 | 6.5 |

In Bangladesh, the prevalence of maternal delivery at home exhibited a marked decline over the past decade, decreasing from 68.1% [66.0-70.0%] in 2012-2013 to 46.4% [45.0-48.0%] in 2019 and 34.9% [32.0-37.0%] in 2022. Regional variation was evident, with Khulna Division demonstrating the lowest prevalence of maternal delivery at home across all years. In contrast, Barishal had the highest prevalence in 2012-13 and 2022, followed by the Sylhet division (**Figure 1**).

**Figure 1:**
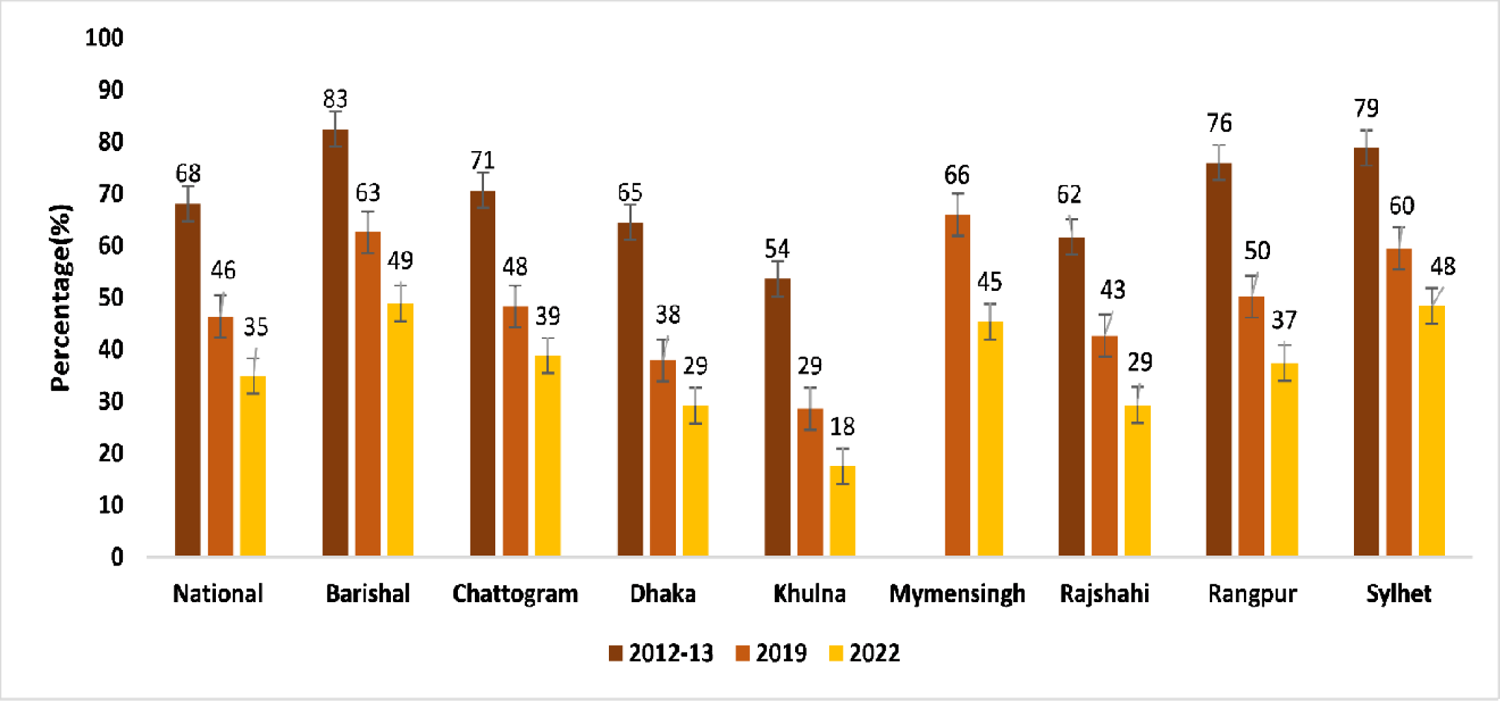
National and divisional estimates of maternal delivery at home in Bangladesh, 2012-2022

Although regional variations were evident, district-level disparities in maternal delivery at home were more pronounced (**Table S1 and** Figure 2). In 2022, the prevalence of maternal delivery at home was >50% in 14 of 64 districts. The highest prevalence was observed in Gopalganj (68.8%), Khagrachhari (68.5%), and Lalmonirhat (66.3%), while the lowest was reported in Bandarban (0%), Chuadanga (0%), and Kushtia (6.5%). In 2012, home-based maternal delivery was most prevalent in hill tract districts such as Bandarban (97.2%) and Rangamati (88.6%), and in haor/coastal districts including Sunamganj (89.3%), Noakhali (88.1%), and Patuakhali (88.0%). By 2019, the burden remained high in hill districts-84.6% in Bandarban, 73.0% in Rangamati, and 76.1% in Khagrachhari. Despite a declining national trend, hill tract and northeastern districts had persistently high prevalence of maternal delivery at home.

**Figure 2:**
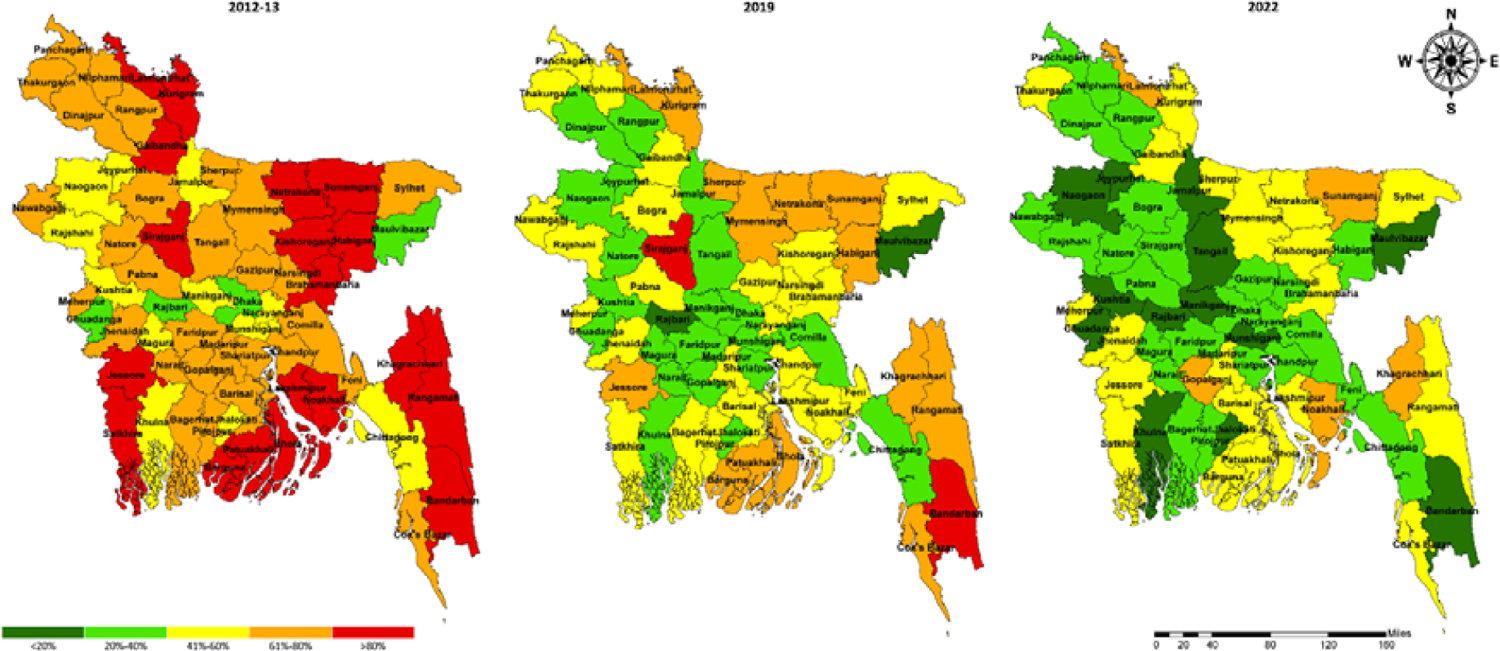
Distribution of maternal delivery at home at the subnational level in Bangladesh, 2012--2022

### Subnational level variations in maternal delivery at home

Despite national and subnational reduction, maternal delivery at home remained common among women aged 35-49 years, although the prevalence in this subgroup declined disproportionately from 79.1% in 2012-2013 to 49.1% in 2022. Women with no formal education consistently showed the highest prevalence of maternal delivery at home (87.5% in 2012-2013, 75.6% in 2019 and 55.7% in 2022), in contrast to women with higher education, whose prevalence fell from 32.2% to 12.6% over the same period. Muslim women persistently reported higher percentage of maternal delivery at home than non-Muslim women (68.9% vs 58.9% in 2012-2013, 47.5% vs 34.1% in 2019 and 36.4% vs 16.1% in 2022). No media exposure was also associated with a higher prevalence of maternal delivery at home, though the prevalence declined notably from 78.3% in 2012 to 51.6% in 2022. Women receiving four or more ANC visits had significantly lower prevalence of maternal delivery at home than those receiving none (38.6% vs 91.1% in 2012, 23.9% vs 80.5% in 2019 and 19.2% vs 76.3% in 2022). Higher parity (3+ children) was associated with increased maternal delivery at home relative to lower parity (1-2 children) over years (81.0% vs 61.8% in 2012, 60.9% vs 39.9% in 2019, and 50.1% vs 28.9% in 2022). Furthermore, women of households headed by individuals with no formal education, those from the poorest wealth quintile, rural residents exhibited consistently higher prevalence of home-based maternal delivery. No significant differences in the prevalence of maternal delivery at home were observed in 2022 based on partner’s current age, spousal age gap or household size (**Table 2**).

**Table 2:**
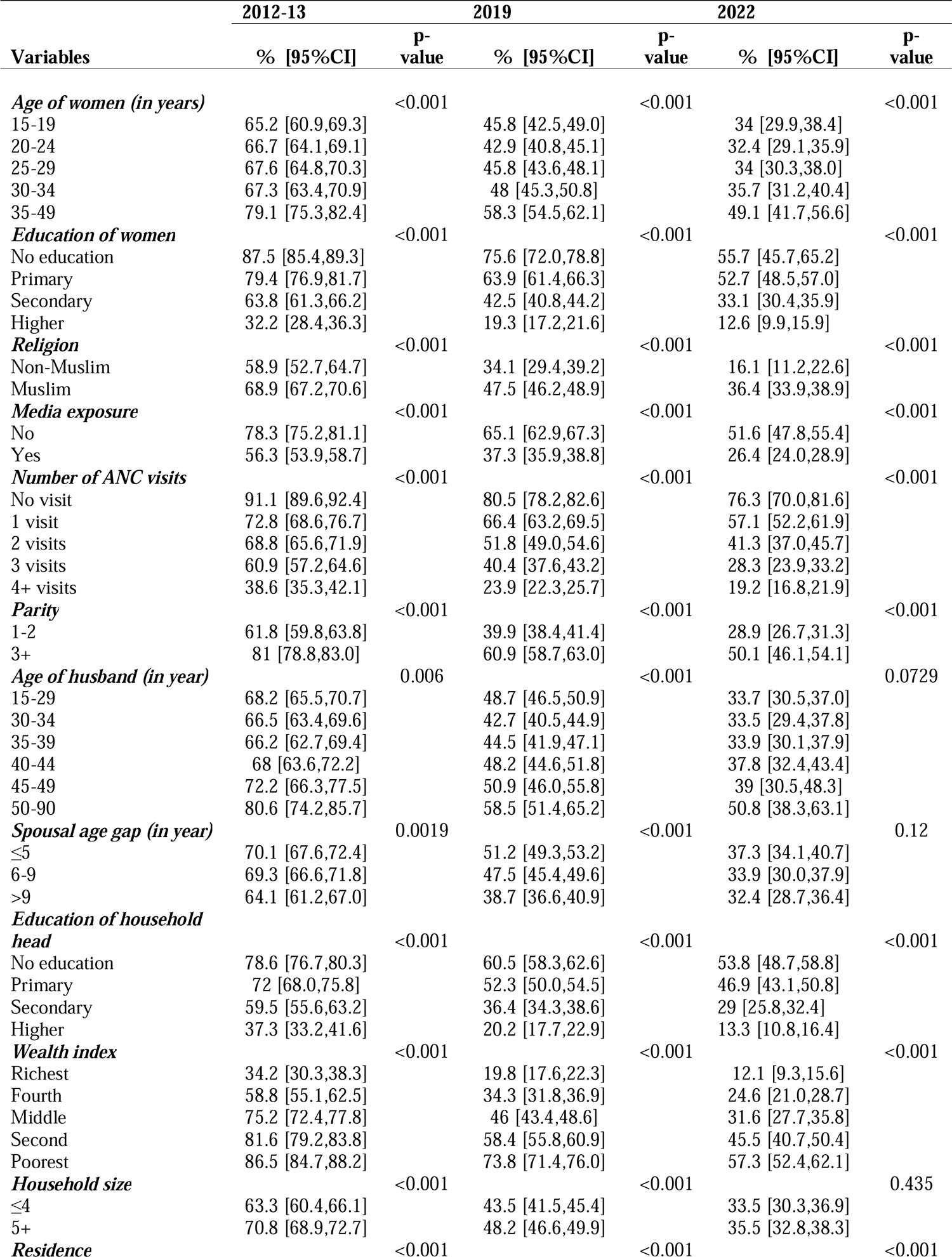
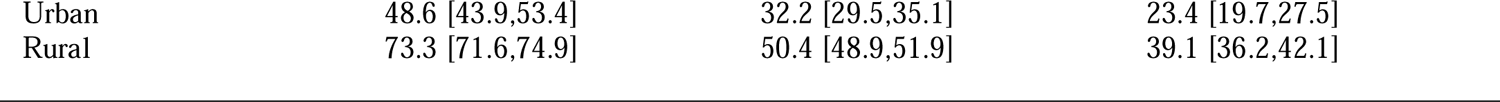
Prevalence (weighted percentage) of maternal delivery at home by sociodemographic characteristics in Bangladesh, 2012-2022.

|  | 2012-13 |  | 2019 |  | 2022 |  |
| --- | --- | --- | --- | --- | --- | --- |
| Variables | % [95%CI] | P-value | % [95%CI] | P-value | % [95%CI] | P-value |
| **Age of women (in years)** |  | <0.001 |  | <0.001 |  | <0.001 |
| 15-19 | 65.2 [60.9,69.3] |  | 45.8 [42.5,49.0] |  | 34 [29.9,38.4] |  |
| 20-24 | 66.7 [64.1,69.1] |  | 42.9 [40.8,45.1] |  | 32.4 [29.1,35.9] |  |
| 25-29 | 67.6 [64.8,70.3] |  | 45.8 [43.6,48.1] |  | 34 [30.3,38.0] |  |
| 30-34 | 67.3 [63.4,70.9] |  | 48 [45.3,50.8] |  | 35.7 [31.2,40.4] |  |
| 35-49 | 79.1 [75.3,82.4] |  | 58.3 [54.5,62.1] |  | 49.1 [41.7,56.6] |  |
| **Education of women** |  | <0.001 |  | <0.001 |  | <0.001 |
| No education | 87.5 [85.4,89.3] |  | 75.6 [72.0,78.8] |  | 55.7 [45.7,65.2] |  |
| Primary | 79.4 [76.9,81.7] |  | 63.9 [61.4,66.3] |  | 52.7 [48.5,57.0] |  |
| Secondary | 63.8 [61.3,66.2] |  | 42.5 [40.8,44.2] |  | 33.1 [30.4,35.9] |  |
| Higher | 32.2 [28.4,36.3] |  | 19.3 [17.2,21.6] |  | 12.6 [9.9,15.9] |  |
| **Religion** |  | <0.001 |  | <0.001 |  | <0.001 |
| Non-Muslim | 58.9 [52.7,64.7] |  | 34.1 [29.4,39.2] |  | 16.1 [11.2,22.6] |  |
| Muslim | 68.9 [67.2,70.6] |  | 47.5 [46.2,48.9] |  | 36.4 [33.9,38.9] |  |
| **Media exposure** |  | <0.001 |  | <0.001 |  | <0.001 |
| No | 78.3 [75.2,81.1] |  | 65.1 [62.9,67.3] |  | 51.6 [47.8,55.4] |  |
| Yes | 56.3 [53.9,58.7] |  | 37.3 [35.9,38.8] |  | 26.4 [24.0,28.9] |  |
| **Number of ANC visits** |  | <0.001 |  | <0.001 |  | <0.001 |
| No visit | 91.1 [89.6,92.4] |  | 80.5 [78.2,82.6] |  | 76.3 [70.0,81.6] |  |
| 1 visit | 72.8 [68.6,76.7] |  | 66.4 [63.2,69.5] |  | 57.1 [52.2,61.9] |  |
| 2 visits | 68.8 [65.6,71.9] |  | 51.8 [49.0,54.6] |  | 41.3 [37.0,45.7] |  |
| 3 visits | 60.9 [57.2,64.6] |  | 40.4 [37.6,43.2] |  | 28.3 [23.9,33.2] |  |
| 4+ visits | 38.6 [35.3,42.1] |  | 23.9 [22.3,25.7] |  | 19.2 [16.8,21.9] |  |
| **Parity** |  | <0.001 |  | <0.001 |  | <0.001 |
| 1-2 | 61.8 [59.8,63.8] |  | 39.9 [38.4,41.4] |  | 28.9 [26.7,31.3] |  |
| 3+ | 81 [78.8,83.0] |  | 60.9 [58.7,63.0] |  | 50.1 [46.1,54.1] |  |
| **Age of husband (in year)** |  | 0.006 |  | <0.001 |  | 0.0729 |
| 15-29 | 68.2 [65.5,70.7] |  | 48.7 [46.5,50.9] |  | 33.7 [30.5,37.0] |  |
| 30-34 | 66.5 [63.4,69.6] |  | 42.7 [40.5,44.9] |  | 33.5 [29.4,37.8] |  |
| 35-39 | 66.2 [62.7,69.4] |  | 44.5 [41.9,47.1] |  | 33.9 [30.1,37.9] |  |
| 40-44 | 68 [63.6,72.2] |  | 48.2 [44.6,51.8] |  | 37.8 [32.4,43.4] |  |
| 45-49 | 72.2 [66.3,77.5] |  | 50.9 [46.0,55.8] |  | 39 [30.5,48.3] |  |
| 50-90 | 80.6 [74.2,85.7] |  | 58.5 [51.4,65.2] |  | 50.8 [38.3,63.1] |  |
| **Spousal age gap (in year)** |  | 0.0019 |  | <0.001 |  | 0.12 |
| ≤5 | 70.1 [67.6,72.4] |  | 51.2 [49.3,53.2] |  | 37.3 [34.1,40.7] |  |
| 6-9 | 69.3 [66.6,71.8] |  | 47.5 [45.4,49.6] |  | 33.9 [30.0,37.9] |  |
| >9 | 64.1 [61.2,67.0] |  | 38.7 [36.6,40.9] |  | 32.4 [28.7,36.4] |  |
| **Education of household head** |  | <0.001 |  | <0.001 |  | <0.001 |
| No education | 78.6 [76.7,80.3] |  | 60.5 [58.3,62.6] |  | 53.8 [48.7,58.8] |  |
| Primary | 72 [68.0,75.8] |  | 52.3 [50.0,54.5] |  | 46.9 [43.1,50.8] |  |
| Secondary | 59.5 [55.6,63.2] |  | 36.4 [34.3,38.6] |  | 29 [25.8,32.4] |  |
| Higher | 37.3 [33.2,41.6] |  | 20.2 [17.7,22.9] |  | 13.3 [10.8,16.4] |  |
| **Wealth index** |  | <0.001 |  | <0.001 |  | <0.001 |
| Richest | 34.2 [30.3,38.3] |  | 19.8 [17.6,22.3] |  | 12.1 [9.3,15.6] |  |
| Fourth | 58.8 [55.1,62.5] |  | 34.3 [31.8,36.9] |  | 24.6 [21.0,28.7] |  |
| Middle | 75.2 [72.4,77.8] |  | 46 [43.4,48.6] |  | 31.6 [27.7,35.8] |  |
| Second | 81.6 [79.2,83.8] |  | 58.4 [55.8,60.9] |  | 45.5 [40.7,50.4] |  |
| Poorest | 86.5 [84.7,88.2] |  | 73.8 [71.4,76.0] |  | 57.3 [52.4,62.1] |  |
| **Household size** |  | <0.001 |  | <0.001 |  | 0.435 |
| ≤4 | 63.3 [60.4,66.1] |  | 43.5 [41.5,45.4] |  | 33.5 [30.3,36.9] |  |
| 5+ | 70.8 [68.9,72.7] |  | 48.2 [46.6,49.9] |  | 35.5 [32.8,38.3] |  |
| **Residence** |  | <0.001 |  | <0.001 |  | <0.001 |

|  |  |  |  |
| --- | --- | --- | --- |
| Urban | 48.6 [43.9,53.4] | 32.2 [29.5,35.1] | 23.4 [19.7,27.5] |
| Rural | 73.3 [71.6,74.9] | 50.4 [48.9,51.9] | 39.1 [36.2,42.1] |

### Determinants of maternal delivery at home

In multivariable analyses, women’s religious affiliation, ANC utilization, parity, household head’s educational attainment, household wealth status, and administrative division were consistently associated with the likelihood of home delivery across survey years. Muslim women exhibited significantly higher odds of maternal delivery at home compared to non-Muslim women, with the association strengthening over time-most notably in 2022 (AOR 3.20, 95% CI 1.99-5.14), relative to 2019 (AOR 1.99, 95% CI 1.60-2.47) and 2012-2013 (AOR 1.36, 95% CI 1.02-1.82). A clear inverse relationship was observed between ANC visit frequency and home delivery; women with four or more ANC visits had substantially lower odds of home delivery compared to those with no ANC visits, with similar magnitudes across all survey years (2012-13: AOR 0.16, 95% CI 0.12-0.20; 2019: AOR 0.17, 95% CI 0.14-0.20; 2022: AOR 0.16, 95% CI 0.11-0.23). Higher parity (≥3 children) was consistently associated with increased odds of maternal delivery at home (2012-13: AOR 1.73, 95% CI 1.25-2.39; 2019: AOR 1.73, 95% CI 1.46-2.05; 2022: AOR 1.81, 95% CI 1.40-2.35). Women residing in households led by individuals with higher educational attainment were significantly less likely to deliver at home compared to those in households headed by individuals with no formal education (2012-13: AOR 0.65, 95% CI 0.50-0.86; 2019: AOR 0.65, 95% CI 0.51-0.82; 2022: AOR 0.50, 95% CI 0.36-0.71). A strong socioeconomic gradient was evident, wherein women from the poorest households had markedly higher odds of maternal delivery at home compared to those from the wealthiest households (2012-13: AOR 2.54, 95% CI 1.80-3.59; 2019: AOR 3.34, 95% CI 2.59-4.30; 2022: AOR 2.84, 95% CI 1.86-4.33). Regionally, women from Khulna division consistently showed lower odds of home delivery (2012-13: AOR 0.61, 95% CI 0.48-0.78; 2019: AOR 0.59, 95% CI 0.47-0.74; 2022: AOR 0.38, 95% CI 0.26-0.56).

Furthermore, women exposed to media were less likely than their counterparts for maternal delivery at home in recent years (2019: AOR 0.78, 95% CI 0.68-0.89; 2022: AOR 0.75, 95% CI 0.61-0.92). In contrast, maternal age, educational attainment, and urban-rural residence were significantly associated with maternal delivery at home in earlier survey rounds (2012– 2013 and 2019), but their effects were less prominent in 2022 (Figure 3 **and Table S2**).

**Figure 3:**
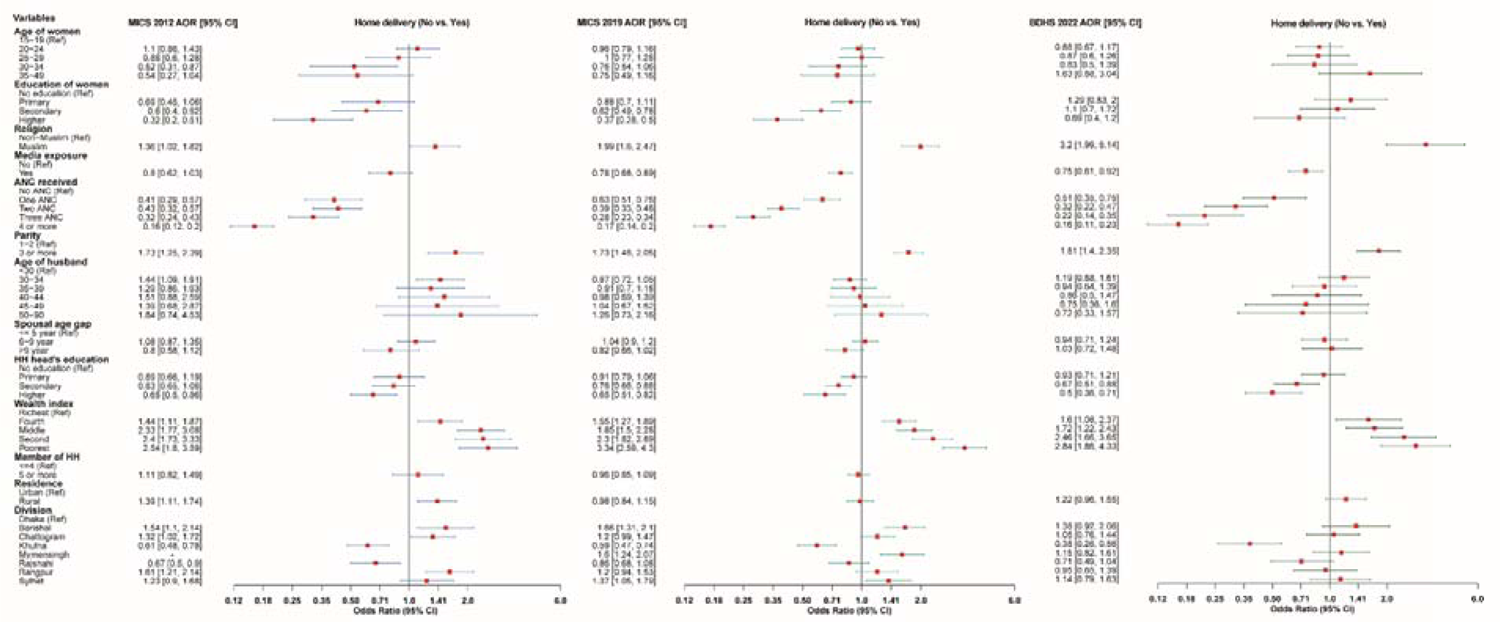
Determinants of maternal delivery at home in Bangladesh, 2012-2022

The frequency of ANC visits and household wealth were consistently associated with maternal delivery at home across both urban and rural settings. Specifically, women who attended four or more ANC visits and those from the poorest households demonstrated a lower likelihood of maternal delivery at home compared to their counterparts. In rural areas, maternal delivery at home was significantly associated over time with religious affiliation, media exposure, parity, and the educational attainment of the household head. However, these associations were not consistently observed among urban women across survey periods (**Table 3**).

**Table 3:** Determinants of maternal delivery at home in Bangladesh by place of residence, 2012-2022.

|  | 2012 | 2019 | 2022 | 2012 | 2019 | 2022 |
| --- | --- | --- | --- | --- | --- | --- |
|  | Urban | Urban | Urban | Rural | Rural | Rural |
| Variables | AOR (95% CI) | AOR (95% CI) | AOR (95% CI) | AOR (95% CI) | AOR (95% CI) | AOR (95% CI) |
| *Age of women* |  |  |  |  |  |  |
| 15-19y | 1 | 1 | 1 | 1 | 1 | 1 |
| 20-24y | 1.51[0.73,3.1] | 1.16 [0.7-1.93] | 1.7[0.88,3.29] | 1.03 [0.79,1.34] | 0.93 [0.76-1.14] | 0.75 [0.55,1.03] |
| 25-29y | 0.8[0.32,1.99] | 1.08 [0.56-2.07] | 0.95 [0.35,2.56] | 1 [0.64,1.55] | 1.02 [0.78-1.34] | 0.86 [0.57,1.29] |
| 30-34y | 0.47[(0.12,1.8] | 0.41 [0.17-1.01] | 1.19 [0.38,3.75] | 0.6 [0.33,1.08] | 0.92 [0.65-1.32] | 0.81 [0.45,1.48] |
| 35-49y | 0.64[0.13,3.02] | 0.66 [0.22-2.13] | 5.63 [1.59,19.92]** | 0.52 [0.24,1.09] | 0.79 [0.5-1.26] | 1.2 [0.58,2.49] |
| *Education of women* |  |  |  |  |  |  |
| No education | 1 | 1 | 1 | 1 | 1 | 1 |
| Primary | 0.18 [0.07,0.46]*** | 1.26 [0.69,2.32] | 1.58 [0.7,3.54] | 1.11 [0.72,1.72] | 0.79 [0.61,1.02] | 1.2[0.71,2.04] |
| Secondary | 0.29 [(0.12,0.7]** | 0.67 [0.38,1.2] | 1.37 [0.57,3.28] | 0.81 [0.53,1.23] | 0.61[0.47,0.79]*** | 1.03[0.6,1.76] |
| Higher | 0.11 [0.04,0.27]*** | 0.3 [0.14,0.64]** | 1.03 [0.34,3.11] | 0.47 [0.29,0.77]** | 0.41[0.3,0.56]*** | 0.65[0.34,1.24] |
| *Religion* |  |  |  |  |  |  |
| Non-muslim |  | 1 | 1 | 1 | 1 | 1 |
| Muslim | 1.34[0.65,2.77] | 1.90 [1.1-3.26]* | 2.16[0.84,5.56] | 1.43[1.02,2.01]* | 1.98 [1.56-2.52]*** | 3.29[1.92,5.64]* |
| *Media exposure* |  |  |  |  |  |  |
| No | 1 | 1 | 1 | 1 | 1 | 1 |
| yes | 1.72[0.83,3.56] | 0.89 [0.62-1.28] | 0.85[0.51,1.43] | 0.71[0.54,0.93]* | 0.78 [0.67-0.88]*** | 0.73[0.59,0.91]** |
| *Number of ANC visits* |  |  |  |  |  |  |
| No visit | 1 | 1 | 1 | 1 | 1 | 1 |
| 1 visit | 0.42[0.18,0.97]* | 0.91 [0.49-1.7] | 0.27[0.11,0.66]** | 0.4[0.27,0.6]*** | 0.59 [0.47-0.75]*** | 0.57[0.37,0.88]* |
| 2 visits | 0.69[0.32,1.5] | 0.38 [0.22-0.65]*** | 0.36[0.15,0.9]* | 0.37[0.28,0.5]*** | 0.39 [0.32-0.48]*** | 0.32[0.21,0.48]*** |
| 3 visits | 0.38[0.19,0.76]** | 0.23 [0.13-0.41]*** | 0.25[0.09,0.68]** | 0.29[0.21,0.4]*** | 0.29 [(0.23-0.35)]*** | 0.22[0.13,0.36]*** |
| 4+ visits | 0.12[0.06,0.24]*** | 0.14 [0.08-0.24]*** | 0.11[0.05,0.27]*** | 0.16[0.12,0.22]*** | 0.18 [0.15-0.22]*** | 0.17[0.11,0.27]*** |
| **Parity** |  |  |  |  |  |  |
| 1-2 | 1 | 1 | 1 | 1 | 1 | 1 |
| 3 or more | 1.1[0.54,2.23] | 1.95 [1.25-3.02]** | 1.37[0.73,2.57] | 1.86[1.22,2.84]** | 1.67 [1.39-2]*** | 2.04[1.49,2.78]*** |
| **Age of husband (in year)** |  |  |  |  |  |  |
| 14-29 | 1 | 1 | 1 | 1 | 1 | 1 |
| 30-34 | 2.59[1.4,4.77]** | 0.71 [0.46-1.11] | 0.76[0.37,1.55] | 1.19[0.88,1.61] | 0.93 [0.76-1.14] | 1.31[0.94,1.82] |
| 35-39 | 2.13[0.89,5.08] | 0.87 [0.46-1.62] | 0.54[0.22,1.32] | 1.06[0.66,1.7] | 0.95 [0.72-1.26] | 0.99[0.64,1.54] |
| 40-44 | 4.59[1.41,14.98]* | 0.87 [0.35-2.16] | 0.68[0.25,1.89] | 1.08[0.56,2.05] | 1.02 [0.7-1.5] | 0.83[0.45,1.53] |
| 45-49 | 1[0.17,5.93] | 1.17 [0.39-3.5] | 0.63[0.15,2.61] | 1.31[0.57,2.99] | 1.01 [0.62-1.65] | 0.64[0.27,1.53] |
| 50-90 | 4.43[0.62,31.86] | 0.56 [0.14-2.22] | 0.22[0.05,1.03] | 1.28[0.46,3.53] | 1.45 [0.79-2.65] | 0.87[0.35,2.12] |
| **Spousal age gap (in year)** |  |  |  |  |  |  |
| <= 5 year | 1 | 1 | 1 | 1 | 1 | 1 |
| 6-9 year | 1.27[0.79,2.02] | 0.93 [0.64-1.34] | 1.07[0.59,1.91] | 1.02[0.8,1.3] | 1.07 [0.91-1.26] | 0.94[0.68,1.3] |
| >9 year | 0.53[0.25,1.12] | 0.73 [0.42-1.25] | 1.59[0.78,3.22] | 0.94[0.65,1.35] | 0.85 [0.67-1.08] | 0.98[0.65,1.48] |
| **Education of household head** |  |  |  |  |  |  |
| No education | 1 | 1 | 1 | 1 | 1 | 1 |
| Primary | 1.05[0.53,2.08] | 0.79[0.53,1.19] | 0.88[0.47,1.65] | 0.83[0.61,1.13] | 0.94[0.8,1.09] | 0.9[0.67,1.2] |
| Secondary | 0.68[0.38,1.23] | 0.64[0.44,0.93]* | 0.56[0.32,0.99]* | 0.85[0.66,1.11] | 0.78[0.67,0.92]** | 0.67[0.5,0.91]** |
| Higher | 0.57[0.31,1.07] | 0.63[0.36,1.1] | 0.35[0.17,0.72]** | 0.68[0.5,0.92]* | 0.66[0.5,0.86]** | 0.54[0.36,0.8]** |
| **Wealth index** |  |  |  |  |  |  |
| Richest | 1 | 1 | 1 | 1 | 1 | 1 |
| Fourth | 1.82[1.07,3.09]* | 1.38 [0.95-2] | 1.95[0.9,4.25] | 1.4[1.07,1.81]* | 1.78 [1.37-2.31]*** | 1.29[0.82,2.03] |
| Middle | 3.17[1.48,6.82]** | 1.39[(0.89-2.17] | 2.83[1.48,5.41]** | 2.31[1.69,3.17]*** | 2.23 [1.71-2.9]*** | 1.22[0.81,1.84] |
| Second | 4.66[1.53,14.22]** | 1.98 [1.17-3.34]* | 4.29[1.96,9.4]*** | 2.18[1.51,3.14]*** | 2.77 [2.09-3.67]*** | 1.82[1.15,2.88]* |
| Poorest | 4.44[1.84,10.74]** | 2.17 [1.13-4.17]* | 6.29[2.52,15.74]*** | 2.35[1.55,3.56]*** | 4.10 [3.04-5.53]*** | 2.11[1.31,3.41]** |
| **Household size** |  |  |  |  |  |  |
| ≤4 | 1 | 1 | 1 | 1 | 1 | 1 |
| 5+ | 1.2[0.73,1.96] | 0.90 [0.66-1.22] | 0.65[0.39,1.07] | 1.06[0.83,1.35] | 1.00 [0.87-1.15] | 0.98[0.78,1.23] |
| **Division** |  |  |  |  |  |  |
| Dhaka | 1 | 1 | 1 | 1 | 1 | 1 |
| Barishal | 0.92[0.44,1.9] | 1.20[0.67,2.15] | 0.49[0.23,1.01] | 1.83[1.26,2.66]** | 1.9[1.45,2.48]*** | 1.56[0.97,2.51] |
| Chattogram | 0.79[0.48,1.31] | 0.96[0.67,1.38] | 1.55[0.9,2.68] | 1.65[1.14,2.38]** | 1.32[1.04,1.68]** | 0.94[0.62,1.42] |
| Khulna | 0.37[0.21,0.67]** | 0.33[0.17,0.64]** | 0.32[0.12,0.9]* | 0.74[0.54,1.01] | 0.68[0.53,0.87]** | 0.39[0.25,0.61]*** |
| Mymensingh |  | 0.42[0.28,0.72]** | 1.33(0.71,2.47] |  | 2.1[1.56,2.69]*** | 1.16[0.78,1.74] |
| Rajshahi | 0.46[0.21,1.03] | 0.60[0.36,0.99]* | 0.44[0.19,1]* | 0.76[0.54,1.07] | 0.93[0.74,1.18] | 0.75[0.49,1.17] |
| Rangpur | 1.77[0.81,3.9] | 1.42[0.77,3.62] | 0.66[0.26,1.65] | 1.67[1.18,2.37]** | 1.24[0.95,1.54] | 1.01[0.65,1.57] |
| Sylhet | 1.05[0.45,2.44] | 1.49[0.85,2.63] | 0.76[0.4,1.47] | 1.37[0.91,2.05] | 1.55[1.17,2.06]** | 1.18[0.77,1.8] |

**Note:** AOR= Adjusted Odds Ratio, CI= Confidence interval, \* p-value<0.05, \*\* p-value<0.01, \*\*\* p-value<0.001

## Discussion

This study provides important insights into the changing landscape and persistent disparities of maternal delivery at home in Bangladesh over the past decade. Although the national prevalence of maternal delivery at home has declined markedly from 68.0% in 2012 to 34.9% in 2022, our findings reveal that this progress is not uniform across the country. Maternal delivery at home varied substantially at the district level, with the hill track districts-such as Rangamati, Bandarban, and Khagrachari-continuing to report rates exceeding 60% in 2022, nearly double the national average. These geographic inequities highlight the limited reach of facility-based delivery interventions in remote and marginalized areas, where barriers such as poor infrastructure, entrenched cultural preferences, and restricted access to healthcare persist despite national improvements.

Our analysis also identified several key factors associated with maternal delivery at home, including religion, number of antenatal care (ANC) visits, parity, education of the household head, household wealth, and administrative division. Muslim women were more likely to have a maternal delivery at home compared to non-Muslims. Women with fewer than four ANC visits, those with higher parity, and those from the poorest households were significantly more likely to experience maternal delivery at home. Importantly, our findings show that the determinants of maternal delivery at home are not consistent between urban and rural areas, suggesting that context-specific, modifiable factors must be considered when designing interventions. While household wealth, ANC visits, and parity were the strongest predictors in rural settings, in urban areas, religion and the education level of the household head played a more prominent role. This urban-rural divergence indicates that interventions must be tailored to the unique drivers of maternal delivery at home in each setting-prioritizing economic empowerment and improved ANC access in rural areas, and focusing on education and cultural engagement in urban contexts.

Our findings reveal substantial division- and district-level differences in home delivery rates. Home delivery remained particularly prevalent in the Barisal, Sylhet, and Mymensingh divisions, where approximately half of the deliveries still occurred at home. These regions have historically faced challenges in healthcare infrastructure, literacy rates, and accessibility, which may contribute to the sustained high prevalence of home deliveries [30]. Previous studies have similarly highlighted that healthcare services in the Mymensingh and Barisal divisions are inadequate, which may further explain the persistence of home deliveries in these areas [31,32]. Similar findings were also reported in different countries, which reported that home delivery rates were typically higher in less developed areas with poor transportation and access to healthcare [33,34]. At the district level, home deliveries were most common in peripheral and remote areas such as Bandarban, Rangamati, Khagrachari, Bhola, and Habiganj. This pattern aligns with findings from Nepal, where home delivery rates were notably higher in mountainous regions due to limited access to healthcare facilities [35].

One important factor influencing home delivery was found to be educational attainment. Compared with women without formal education, those with greater levels of education had a much lower likelihood of giving birth at home. This result is in line with other research from several nations, indicating that education is crucial in influencing maternal healthcare choices [36,37]. Awareness and knowledge might be the main reasons for this type of scenario, as women with lower levels of education may possess limited awareness regarding maternal healthcare facilities and the advantages of in-facility deliveries [37–39]. Education empowers women to efficiently navigate healthcare institutions and make informed decisions about their health [37]. While the gap in home delivery rates between educated and less educated women has narrowed over time, further efforts are required to ensure universal access to facility-based deliveries, particularly among disadvantaged groups.

Home delivery rates are strongly impacted by the household head’s educational attainment in addition to the mother’s education. The more educated the household head was, the less likely the woman was to give birth at home. According to a Nigerian study, households with educated heads spent more money on healthcare, which increased the rates of institutional delivery [40]. Another study in Bangladesh revealed that a lower education level of household heads decreases maternal healthcare utilization in Bangladesh [41]. These findings emphasize the broader role of education not only in individual decision-making but also in shaping household-level health-seeking behaviors.

Media exposure was identified as a protective factor against home delivery. Increasing exposure to media can alter perceptions positively and encourage expectant mothers to choose facility-based deliveries over home deliveries, as public awareness and health-related behaviors are strongly influenced by media exposure. Owing to a lack of knowledge, cultural customs, and restricted access to medical facilities, home deliveries continue in many areas. To promote institutional deliveries and enhance the health of mothers and newborns, increasing media exposure can be a useful tactic [42,43]. In Nigeria, similar findings have been reported, where increased usage of maternal healthcare services was linked to media exposure [44]. Expanding media-based health awareness programs could be an effective strategy to reduce home delivery rates further.

Religious beliefs also play a role in home delivery choices. Our research suggests that Muslim women exhibited a greater tendency for home delivery than their non-Muslim peers did. Cultural norms, traditional beliefs, and socioeconomic factors may contribute to this trend [45]. A similar pattern has been observed in India, where women of the Muslim faith exhibited a greater propensity for home births [36]. However, further research is necessary to investigate the underlying causes of this difference and to develop culturally relevant treatments that encourage institutional deliveries within various religious communities.

The utilization of antenatal care (ANC) is a crucial factor influencing home delivery, as women who visit four or more ANC appointments are much less inclined to deliver at home. This aligns with findings from Afghanistan, where ANC visits were strongly associated with lower home deliveries [46]. High-quality ANC services provide essential counseling, health screenings, and education on the benefits of skilled birth attendance, which collectively influence women’s decisions to seek facility-based delivery over home delivery [47–51]. The consistent relationship between ANC utilization and a lower likelihood of home delivery highlights the need to strengthen ANC services and ensure equitable access for all pregnant women.

Parity, or the number of previous childbirths, also influenced home delivery rates. Home deliveries were more common among women with three or more children. This finding is similar to those of studies conducted in Bangladesh and Ethiopia, which reported that women who are first-time mothers or have fewer pregnancies are more likely to choose facility-based births [30,37]. Likewise, multiparous women in rural Zambia are more inclined to give birth at home [52]. Women with multiple previous births may feel more confident about delivering at home, particularly if they have had uncomplicated deliveries in the past. Furthermore, multiparous women may be more likely to opt for delivery at home because of logistical difficulties, financial limitations, and a dependence on traditional birth attendants. Addressing these barriers through community-based interventions and financial support programs could help encourage facility-based deliveries among high-parity women.

Our findings highlight a strong wealth disparity in home delivery rates. Women from poorer households were significantly more likely to give birth at home than those from affluent households were. Similar findings have been reported in several countries, including Nigeria, Nepal, and India [53–56]. Financial constraints, out-of-pocket fees, and indirect costs such as transportation and lost wages may deter women from seeking facility-based deliveries. Studies from Kenya and West Africa similarly report persistent economic disparities in maternal healthcare utilization, emphasizing the need for financial support mechanisms such as conditional cash transfers and subsidies to promote institutional deliveries among economically disadvantaged groups [47,48,57].

Urban rural differences in home delivery rates were also evident. Home deliveries were more common among women in rural areas. According to some studies conducted in Africa and Indonesia, urban women are significantly more likely than their rural counterparts are to give birth in a medical facility [58–60]. Women living in rural areas confront unique obstacles to facility delivery, such as a lack of qualified healthcare personnel, restricted availability of medical services, and poor healthcare infrastructure [44,49,61,62]. Bridging the urban rural gap in maternal healthcare requires strategic investments in rural healthcare facilities, improved transportation networks, and community-based healthcare interventions.

Bangladesh has implemented a range of interventions over the past decade-including the expansion of community clinics, maternal health voucher schemes, and awareness campaigns-that have contributed to the overall reduction in maternal delivery at home and increased skilled birth attendance [63]. The Maternal Health Voucher Scheme (MHVS), launched in 2007, has been particularly impactful in improving access to antenatal care, skilled assistance at delivery, and postnatal care among disadvantaged women, though administrative and coverage challenges remain [64,65]. Our study affirms the success of these efforts at the national level but also reveals persistent gaps: maternal delivery at home remains disproportionately high among marginalized groups such as women in the hill tracts, high-parity mothers, and the poorest households. Additionally, our findings highlight that determinants of maternal delivery at home differ between urban and rural settings, with factors like religion and household head’s education being more influential in urban areas, while wealth, parity, and ANC visits are more critical in rural regions. These context-specific disparities suggest that, despite overall progress, current interventions have not fully addressed the unique barriers faced by these populations. To further reduce maternal delivery at home, particularly in high-prevalence areas, our study recommends mobile and culturally sensitive outreach in remote districts, greater engagement of religious and community leaders, incentivization of complete ANC utilization, and adaptation of policies to the distinct socio-demographic realities of different regions.

## Strengths and Limitations

A key strength of this study is its ability to provide valuable insights at both the national and subnational levels using data from the MICS 2012–13 and 2019, as well as the BDHS 2022. The use of the most recent BDHS 2022 dataset, which employed rigorous sampling techniques, enhances the generalizability of the findings to women of reproductive age (15–49 years) across the country. Additionally, the large sample size and application of appropriate statistical techniques allowed for robust identification of significant predictors and assessment of inequality gradients across multiple subgroups. The findings can inform policies aimed at reducing maternal delivery at home clusters and identifying areas with limited access to facility-based childbirth services.

However, the study has certain limitations. First, as with most MICS and DHS-based studies, data are self-reported and thus subject to recall bias and social desirability bias, which may affect the accuracy of reported reproductive behaviors. Second, due to the cross-sectional nature of the survey, the study cannot establish causal relationships between maternal delivery at home and its determinants. Third, some relevant covariates, such as maternal comorbidities, nutritional status, dietary patterns, physical activity, and psychosocial factors, were not included in the BDHS and thus could not be examined, even though they may have significant implications for maternal delivery at home. Fourth, socioeconomic characteristics were measured at the time of the interview, which may not accurately reflect the woman’s status at the time of pregnancy or childbirth. And finally, although the BDHS 2022 incorporated district-level estimates for the first time, the limited sample size in several districts has resulted in unstable prevalence rates, with some districts reporting values of ‘0,’ which is inconsistent with prior nationally representative surveys such as MICS.

## Conclusion

Over the last decade, Bangladesh has made great strides in lowering the country’s home delivery rates. However, one-third of births still occur at home, highlighting ongoing challenges in ensuring universal access to institutional deliveries. Geographic, socioeconomic, and demographic disparities persist, underscoring the need for targeted interventions. Strengthening healthcare infrastructure, particularly in remote and hilly areas, improving maternal health education, and increasing ANC utilization can further reduce home deliveries. Additionally, addressing financial barriers through subsidies and incentive programs may encourage greater use of facility-based care. Culturally sensitive approaches, media-based awareness campaigns, and community engagement initiatives should be prioritized to achieve equitable maternal healthcare access. To develop inclusive and successful policies, future studies should look at how religious, cultural, and economic issues affect decisions about maternal delivery at home. Bangladesh may achieve universal facility-based birthing by resolving these inequities, which would ultimately improve the health of expectant mothers and newborns.

## Data Availability

All data relevant to the study are included in the article or uploaded as supplemental information. All relevant data are within the manuscript. Raw MICS (https://mics.unicef.org/) and BDHS (https://dhsprogram.com/) data are publicly available that can be accessed upon request.

## Declarations

## Acknowledgements

We are grateful to the MICS and DHS team for allowing us to conduct the analysis of this study using the MICS 2012 and 2019 datasets and the BDHS 2022 dataset.

## Author Contributions

Conceptualization, R.B.I; Data curation, R.B.I, and M.L.K; Formal analysis, R.B.I, and M.L.K; Methodology, R.B.I, and S.T.A.N; Visualization, R.B.I, and S.T.A.N; Investigation, S.M., and A.A; Supervision, S.T.A.N; Validation, M.B.L; Writing—original draft, R.B.I, and M.L.K; Writing - review and editing, S.M., A.A., S.T.A.N and M.B.L; All authors have read and agreed to the published version of the manuscript.

## Funding

The authors received no specific grant from funding agencies in the public, commercial, or not-for-profit sectors.

## Preprint

Available on medRxiv.

## Consent for publication

Not applicable.

## Competing interests

There are no potential conflicts (financial, professional, or personal) for any of the authors to disclose.

## Notes

### Competing Interest Statement

The authors have declared no competing interest.

### Funding Statement

The author(s) received no specific funding for this work.

### Author Declarations

This study utilized deidentified secondary data obtained from the UNICEF (https://mics.unicef.org/surveys) and DHS (https://www.dhsprogram.com/data/available-datasets.cfm) websites. Since the data were publicly available and did not contain any personally identifiable information, ethical approval was not needed.

### Summary of Updates

The word "Spatiotemporal" is removed from the manuscript as it was not necessary in the manuscript.

## References

1. Trends in maternal mortality 2000 to 2023: estimates by WHO, UNICEF, UNFPA, World Bank Group and UNDESA/Population Division. [cited 13 May 2025]. Available: https://www.who.int/publications/i/item/9789240108462

2. El-Saharty S, Ohno N. South Asia’s quest for reduced maternal mortality: What the data show. In: World Bank Blog: Investing in Health. 2015.

3. Hossain AT, Siddique AB, Jabeen S, Khan S, Haider MM, Ameen S, et al. Maternal mortality in Bangladesh: Who, when, why, and where? A national survey-based analysis. J Glob Health. 2023;13. doi:10.7189/JOGH.13.07002

4. The DHS Program - Bangladesh: DHS, 2022 - Final Report (English). [cited 13 May 2025]. Available: https://www.dhsprogram.com/publications/publication-fr386-dhs-final-reports.cfm?cssearch=1551986_1

5. Document Viewer. [cited 13 May 2025]. Available: https://docs.un.org/en/A/RES/70/1

6. UNISEF. Maternal and newborn health. 2023.

7. Kitila SB, Feyissa GT, Olika AK, Wordofa MA. Maternal Healthcare in Low- and Middle-Income Countries: A Scoping Review. Health Serv Insights. 2022;15. doi:10.1177/11786329221100310

8. Oladipo IA, Akinwaare MO. Trends and patterns of maternal deaths from 2015 to 2019, associated factors and pregnancy outcomes in rural Lagos, Nigeria: a cross-sectional study. Pan African Medical Journal. 2023;44.

9. Hernández-Vásquez A, Chacón-Torrico H, Bendezu-Quispe G. Prevalence of home birth among 880,345 women in 67 low- and middle-income countries: A meta-analysis of Demographic and Health Surveys. SSM Popul Health. 2021;16. doi:10.1016/j.ssmph.2021.100955

10. Shrestha SK, Banu B, Khanom K, Ali L, Thapa N, Stray-Pedersen B, et al. Changing trends on the place of delivery: Why do Nepali women give birth at home? Reprod Health. 2012;9: 1–8. doi:10.1186/1742-4755-9-25

11. Sahoo J, Singh SV, Gupta VK, Garg S, Kishore J. Do socio-demographic factors still predict the choice of place of delivery: A cross-sectional study in rural North India. J Epidemiol Glob Health. 2015;5: S27–S34.

12. Shrestha SK, Banu B, Khanom K, Ali L, Thapa N, Stray-Pedersen B, et al. Changing trends on the place of delivery: Why do Nepali women give birth at home? Reprod Health. 2012;9: 1–8. doi:10.1186/1742-4755-9-25

13. Tiruneh SA, Lakew AM, Yigizaw ST, Sisay MM, Tessema ZT. Trends and determinants of home delivery in Ethiopia: Further multivariate decomposition analysis of 2005-2016 Ethiopian Demographic Health Surveys. BMJ Open. 2020;10: 1–10. doi:10.1136/bmjopen-2019-034786

14. Caulfield T, Onyo P, Byrne A, Nduba J, Nyagero J, Morgan A, et al. Factors influencing place of delivery for pastoralist women in Kenya: A qualitative study. BMC Womens Health. 2016;16: 1–11. doi:10.1186/s12905-016-0333-3

15. Sarker BK, Rahman M, Rahman T, Hossain J, Reichenbach L, Mitra DK. Reasons for preference of home delivery with traditional birth attendants (TBAs) in Rural Bangladesh: A qualitative exploration. PLoS One. 2016;11. doi:10.1371/journal.pone.0146161

16. Islam MM, Shahjahan M. Exploring the reasons and factors influencing the choice of home delivery of births in rural Bangladesh: a community-based cross-sectional study. J Health Res. 2022;36. doi:10.1108/JHR-07-2020-0284

17. World Health Organization. WORLD HEALTH STATISTICS - MONITORING HEALTH FOR THE SDGs. World Health Organization. 2016; 1.121.

18. Dey R, Dey SR, Haque M, Rahman AB, Kundu S, Setu SP, et al. Mapping the prevalence and covariates associated with home delivery in Bangladesh: A multilevel regression analysis. PLoS One. 2024;19: e0313606.

19. Talukder A, Anik BH, Hossain MI, Haq I, Habib MJ. Socioeconomic and demographic factors for mothers’ delivery at home: a comparative study among BDHS 2007, 2011 and 2014. Asian Journal of Social Health and Behavior. 2022;5: 10–17.

20. Ahmed KT, Karimuzzaman M, Mahmud S, Rahman L, Hossain MM, Rahman A. Influencing factors associated with maternal delivery at home in urban areas: a cross-sectional analysis of the Bangladesh Demographic and Health Survey 2017–2018 data. J Health Popul Nutr. 2023;42: 83.

21. BANGLADESH 2012-13 MICS FINAL REPORT RELEASED | UNICEF MICS. [cited 13 May 2025]. Available: https://mics.unicef.org/news/bangladesh-2012-13-mics-final-report-released

22. Bangladesh 2019 MICS Report - English | PDF | Drinking Water | Sanitation. [cited 13 May 2025]. Available: https://www.scribd.com/document/537704949/Bangladesh-2019-MICS-Report-English

23. Pathey P. Bangladesh multiple indicator cluster survey 2012–2013 Key findings. Bangladesh Bur Stat UNICEF Bangladesh. 2014;2014: 2014.

24. Noor STA, Shil P, Talukdar A, Aktar S, Uddin MJ. Exploring Factors Influencing Wealth-Related Disparities in Institutional Delivery: A Decomposition Analysis Using Bangladesh Multiple Indicator Cluster Survey (MICS) 2019. Public Health Challenges. 2025;4: e70066.

25. National Institute of Population Research and Training (NIPORT), ICF. Bangladesh Demographic and Health Survey 2022 Final Report. 2024. Available: https://www.dhsprogram.com/publications/publication-FR386-DHS-Final-Reports.cfm

26. Ahmed KT, Karimuzzaman Md, Mahmud S, Rahman L, Hossain MdM, Rahman A. Influencing factors associated with maternal delivery at home in urban areas: a cross-sectional analysis of the Bangladesh Demographic and Health Survey 2017–2018 data. J Health Popul Nutr. 2023;42: 83. doi:10.1186/s41043-023-00428-9

27. Enuameh YAK, Okawa S, Asante KP, Kikuchi K, Mahama E, Ansah E, et al. Factors influencing health facility delivery in predominantly rural communities across the three ecological zones in Ghana: A cross-sectional study. PLoS One. 2016;11: 1–16. doi:10.1371/journal.pone.0152235

28. Dey D, Haque MdS, Islam MdM, Aishi UI, Shammy SS, Mayen MdSA, et al. The proper application of logistic regression model in complex survey data: a systematic review. BMC Med Res Methodol. 2025;25: 15. doi:10.1186/s12874-024-02454-5

29. von Elm E, Altman DG, Egger M, Pocock SJ, Gøtzsche PC, Vandenbroucke JP. The Strengthening the Reporting of Observational Studies in Epidemiology (STROBE) Statement: Guidelines for reporting observational studies. International Journal of Surgery. 2014;12: 1495–1499. 10.1016/j.ijsu.2014.07.013

30. Ahmed KT, Karimuzzaman Md, Mahmud S, Rahman L, Hossain MdM, Rahman A. Influencing factors associated with maternal delivery at home in urban areas: a cross-sectional analysis of the Bangladesh Demographic and Health Survey 2017–2018 data. J Health Popul Nutr. 2023;42: 83. doi:10.1186/s41043-023-00428-9

31. Huda TM, Chowdhury M, El Arifeen S, Dibley MJ. Individual and community level factors associated with health facility delivery: A cross sectional multilevel analysis in Bangladesh. PLoS One. 2019;14: e0211113.

32. Badiuzzaman M, Murshed SM, Rieger M. Improving maternal health care in a post conflict setting: evidence from Chittagong Hill tracts of Bangladesh. J Dev Stud. 2020;56: 384–400.

33. Teshale AB, Alem AZ, Yeshaw Y, Kebede SA, Liyew AM, Tesema GA, et al. Exploring spatial variations and factors associated with skilled birth attendant delivery in Ethiopia: geographically weighted regression and multilevel analysis. BMC Public Health. 2020;20: 1–19.

34. Moshi FV, Lymo G, Gibore NS, Kibusi SM. Prevalence and factors associated with home childbirth with unskilled birth assistance in Dodoma-Tanzania: a cross sectional study. East Afr Health Res J. 2020;4: 92.

35. Shahabuddin ASM, De Brouwere V, Adhikari R, Delamou A, Bardaj A, Delvaux T. Determinants of institutional delivery among young married women in Nepal: Evidence from the Nepal Demographic and Health Survey, 2011. BMJ Open. 2017;7: e012446.

36. Sahoo J, Singh SV, Gupta VK, Garg S, Kishore J. Do socio-demographic factors still predict the choice of place of delivery: A cross-sectional study in rural North India. J Epidemiol Glob Health. 2015;5. doi:10.1016/j.jegh.2015.05.002

37. Tiruneh SA, Lakew AM, Yigizaw ST, Sisay MM, Tessema ZT. Trends and determinants of home delivery in Ethiopia: Further multivariate decomposition analysis of 2005-2016 Ethiopian Demographic Health Surveys. BMJ Open. 2020;10: 1–10. doi:10.1136/bmjopen-2019-034786

38. Straneo M, Fogliati P, Azzimonti G, Mangi S, Kisika F. Where do the rural poor deliver when high coverage of health facility delivery is achieved? Findings from a community and hospital survey in Tanzania. PLoS One. 2014;9: 1–17. doi:10.1371/journal.pone.0113995

39. Kruk ME, Hermosilla S, Larson E, Vail D, Chen Q, Mazuguni F, et al. Who is left behind on the road to universal facility delivery? A cross-sectional multilevel analysis in rural Tanzania. Tropical Medicine and International Health. 2015;20: 1057–1066. doi:10.1111/tmi.12518

40. Ogundari K, Abdulai A. Determinants of household’s education and healthcare spending in Nigeria: Evidence from survey data. African Development Review. 2014;26: 1–14.

41. Rahman MM, Amin MT, Ferdous Z, Patwary H, Haider MM. Education of household head and maternal healthcare utilization: the case of Bangladesh. BMC Public Health. 2024;24: 3439.

42. Mills S, Williams JE, Adjuik M, Hodgson A. Use of health professionals for delivery following the availability of free obstetric care in Northern Ghana. Matern Child Health J. 2008;12: 509–518. doi:10.1007/s10995-007-0288-y

43. Agha S, Carton TW. Determinants of institutional delivery in rural Jhang, Pakistan. Int J Equity Health. 2011;10: 1–12. doi:10.1186/1475-9276-10-31

44. Babalola S, Fatusi A. Determinants of use of maternal health services in Nigeria - Looking beyond individual and household factors. BMC Pregnancy Childbirth. 2009;9: 43. doi:10.1186/1471-2393-9-43

45. Kamal SMM, Hassan CH, Alam GM. Determinants of institutional delivery among women in Bangladesh. Asia Pac J Public Health. 2015;27: NP1372–NP1388. doi:10.1177/1010539513486178

46. Rahman M, Saha P, Uddin J. Associations of antenatal care visit with utilization of institutional delivery care services in Afghanistan: intersections of education, wealth, and household decision-making autonomy. BMC Pregnancy Childbirth. 2022;22: 255.

47. Colombara D V., Hernández B, Schaefer A, Zyznieuski N, Bryant MF, Desai SS, et al. Institutional delivery and satisfaction among indigenous and poor women in Guatemala, Mexico, and Panama. PLoS One. 2016;11: 1–17. doi:10.1371/journal.pone.0154388

48. Tekelab T, Yadecha B, Melka AS. Antenatal care and women’s decision making power as determinants of institutional delivery in rural area of Western Ethiopia. BMC Res Notes. 2015;8: 1–8. doi:10.1186/s13104-015-1708-5

49. Séraphin MN, Ngnie-Teta I, Ayoya MA, Khan MR, Striley CW, Boldon E, et al. Determinants of Institutional Delivery Among Women of Childbearing Age in Rural Haiti. Matern Child Health J. 2015;19: 1400–1407. doi:10.1007/s10995-014-1646-1

50. Agha S, Williams E. Quality of antenatal care and household wealth as determinants of institutional delivery in Pakistan: Results of a cross-sectional household survey. Reprod Health. 2016;13: 1–8. doi:10.1186/s12978-016-0201-5

51. Feyissa TR, Genemo GA. Determinants of institutional delivery among childbearing age women in Western Ethiopia, 2013: Unmatched case control study. PLoS One. 2014;9: 1–7. doi:10.1371/journal.pone.0097194

52. Scott NA, Henry EG, Kaiser JL, Mataka K, Rockers PC, Fong RM, et al. Factors affecting home delivery among women living in remote areas of rural Zambia: a cross-sectional, mixed-methods analysis. Int J Womens Health. 2018; 589–601.

53. Newell R, Spillman I, Newell M-L. The use of facilities for labor and delivery: the views of women in rural Uganda. J Public Health Afr. 2017;8: 592.

54. Olorunsaiye CZ, Huber LB, Laditka SB, Kulkarni S, Boyd AS. Factors associated with health facility delivery in West and Central Africa: a multilevel analysis of seven countries. Health Care Women Int. 2020;41: 3–21.

55. Singh PK, Rai RK, Singh L. Examining the effect of household wealth and migration status on safe delivery care in urban India, 1992–2006. 2012.

56. Devkota B, Maskey J, Pandey AR, Karki D, Godwin P, Gartoulla P, et al. Determinants of home delivery in Nepal–A disaggregated analysis of marginalised and non-marginalised women from the 2016 Nepal Demographic and Health Survey. PLoS One. 2020;15: e0228440.

57. Kinuthia J, Kohler P, Okanda J, Otieno G, Odhiambo F, John-Stewart G. A community-based assessment of correlates of facility delivery among HIV-infected women in western Kenya. BMC Pregnancy Childbirth. 2015;15: 1–9. doi:10.1186/s12884-015-0467-6

58. Idris H, Budiastuti A, Razak R, Hasyim H. Delivery Services Utilization Based on Urban Rural Status in Indonesia. Open Access Maced J Med Sci. 2022;10: 1147–1152.

59. Newell R, Spillman I, Newell M-L. The use of facilities for labor and delivery: the views of women in rural Uganda. J Public Health Afr. 2017;8: 592.

60. Moyer CA, Mustafa A. Drivers and deterrents of facility delivery in sub-Saharan Africa: a systematic review. Reprod Health. 2013;10: 1–14.

61. Dahiru T, Oche OM. Determinants of antenatal care, institutional delivery and postnatal care services utilization in Nigeria. Pan African Medical Journal. 2015;21: 1–17. doi:10.11604/pamj.2015.21.321.6527

62. Amano A, Gebeyehu A, Birhanu Z. Institutional delivery service utilization in Munisa Woreda, South East Ethiopia: a community based cross-sectional study. BMC Pregnancy Childbirth. 2012;12: 1–6. doi:10.1186/1471-2393-12-105

63. Mahmood SS, Amos M, Hoque S, Mia MN, Chowdhury AH, Hanifi SMA, et al. Does healthcare voucher provision improve utilisation in the continuum of maternal care for poor pregnant women? Experience from Bangladesh. Glob Health Action. 2019;12. doi:10.1080/16549716.2019.1701324,

64. Chowdhury AH, Hanifi SMA, Iqbal M, Hossain A, Stones W, Amos M, et al. Does maternal health voucher scheme have association with distance inequality in maternal and newborn care utilization? Evidence from rural Bangladesh. PLoS One. 2023;18: e0295306.

65. Talukder MN, Rob U, Musa SAJM, Bajracharya A, Keya KT, Noor FR, et al. Evaluation of the impact of the voucher program for improving maternal health behavior and status in Bangladesh. 2014.

